# The ACUte effects of SITting time on physiological and psychological function in older adults (ACUSIT): An observational cohort study

**DOI:** 10.64898/2026.09.08.26362486

**Authors:** Sandra Agyapong-Badu, Carolyn Greig, Emma Bostock, Kirsty C McGee, Julie Black, Elisa Poses-Ferrer, Thomas Jackson, Carly Welch, Sebastien FM Chastin, Anna C Whittaker, Dawn A Skelton

## Abstract

**Introduction:** There is accumulating evidence supporting the longer-term detrimental health effects of prolonged bouts of sedentary behaviour (sitting). However, there are less data relating to older adults and in particular, the acute dose-response effects of uninterrupted sitting are lacking. This study aimed to identify key physiological and psychological outcomes influenced by acute periods of inactivity.

**Methods:** The study utilised a randomised cross-over design. Participants randomly completed sitting times of 1, 2 and 4 hours on separate days at the same time of day, and were permitted to watch television, knit or read but not to sleep. The sitting times corresponded to a brief control period of sitting (1 hour (h)), moderately sedentary (2-4 h per day) and sedentary (>4 h per day). The primary outcome measure was lower limb power output using the Nottingham Power Rig (Watts (W)/kg). Secondary outcomes included a range of physiological and psychological measures. Assessments were made before and after each period of sitting time in a temperature-controlled room. Differences between the pre and post measurements across each session were analysed using repeated measures ANOVA with post-hoc comparisons to determine between condition differences.

**Results:** Lower limb power output did not change significantly (p = 0.67) following the 1 h (pre: 1.44 ± 0.6, post: 1.461 ± 0.6 W/kg), 2 h (pre 1.42 ± 0.7, post: 1.44 ± 0.6 W/kg) and 4 h (pre: 1.47 ± 0.7, post: 1.44 ± 0.6 W/kg) sitting times. A significant increase in mean systolic and diastolic blood pressure was recorded following the 2h (systolic pre: 137 ± 13, post: 141 ± 6 mmHg; p<0.001; diastolic pre: 74 ± 10, post 76 ± 10mmHg; p<0.001) and 4 h sitting times (systolic pre 137 ± 14, post 147 ± 15 mmHg; p<0.001; diastolic pre 73 ± 10, post 79 ± 10 mmHg; p<0.001). Mean arterial pressure also significantly increased after the 2 h (pre: 95 ± 10, post: 97 ± 11 mmHg; p<0.001) and 4 h (pre: 95 ± 10, post: 102 ± 10 mmHg; p<0.001) periods of sitting. Timed Up & Go performance declined after all sessions of sitting (p=0.01) whereas Digit Symbol Substitution (DSS) scores (p=0.001) and % DSS correct scores (p<0.001) improved. Vitality declined across all sessions, but this was significantly worse after 4 h. Psychological outcome measure subscales (Profile of Mood State; POMS) related to vigour and friendliness showed significant adverse changes across all sessions, although vigour was worst after 4 h sitting. Further, fatigue changes were greater and negative after 2 and 4 h sitting compared to 1 h, and post-sitting values were highest after 4 h. Data obtained for inflammatory markers was inconclusive, due mainly to concentrations being out of range of measurement. In a small subset of participants (n=13), significant increases in bladder volume comparing post with pre-sitting time were found across all 3 sitting periods (p<0.05), however there were no significant differences between sessions.

**Conclusions:** These data show that sitting for 2 h or more may have potentially deleterious effects on cardiovascular biomarkers including blood pressure, as well as increasing the risk of falling. Vitality, vigour and fatigue, subscales of mood may also be adversely affected in older adults. These effects appear to increase with continuous sitting time.

## Introduction

Sedentary behaviour is “any waking behaviour characterized by an energy expenditure ≤1.5 metabolic equivalents (METs), while in a sitting, reclining or lying posture” [1]. A systematic review identifying more than 500,000 older adults (aged 60 years and older) in 19 studies, highlighted a high prevalence of sedentary behaviour (i.e., sitting time), whether quantified indirectly using self-report or directly by accelerometer [2]. Almost 60% of older adults reported sitting for more than 4 hours (h) per day, (consistent with recent global estimates for high income countries; [3]), 65% sat in front of a screen for more than 3 h daily and more than 55% reported watching more than 2 h of TV per day [2], with another review including objective measures reporting that older adults are sedentary 65-80% of their waking day [4]. Other studies report even higher proportions of sedentariness; up to 75% of total accelerometer wear time in community-dwelling older adults [5] and 90% in care home residents [6].

There is substantial evidence from both epidemiological and observational studies that sedentary behaviour is a major modifiable risk factor for chronic disease and predicts cardiovascular and all-cause mortality, independent of the amount of physical activity (PA), [6,7,8,9]. In addition, a dose response relationship has been shown between sitting time and cardiovascular disease and all-cause mortality across the adult age range [10]. A previous review of the physiological and health impacts of sedentary behaviour in older adults reported negative associations (independent of PA), between sedentariness and cardiometabolic and musculoskeletal health [11], with others reporting associations between sedentariness and cognition (executive functioning), [12,13] and sedentariness and quality of life [14]. A systematic review across the adult age range reported insufficient evidence for an association between sitting time and stress although over half of the included studies were reported as being methodologically weak and only 7 included objective measures of stress (i.e., cortisol) [15]. A more recent review, focussing on young adults, found significant positive associations between acute biological stress responses and sedentary behaviour, but less evidence relating to chronic psychological responses, acute psychological stress, and chronic biological stress [16].

The physiological mechanisms underlying changes in physical and psychological health induced by sedentary behaviour in older adults, are relatively under-researched. Limited evidence for non-age specific cardiometabolic, (e.g., increases in pro-inflammatory cytokines, reduced insulin sensitivity), vascular (e.g., increases in blood pressure, endothelial cell damage), and musculoskeletal mechanisms (e.g., reduced bone mineral content), have been proposed [11]. However, knowledge gaps remain with respect to how these mechanisms influence health outcomes or specifically in older adults. A first step to address the existing knowledge gaps would be to study the acute (or ‘single bout’) effects of sedentariness on physiological and psychological health. Previous studies in young adults have reported physiological impairments such as reduced endothelial function after 3h sitting time [17] and a reduction in insulin action after 1 day of sitting [18]. Further studies in middle-aged overweight or obese adults have shown altered cardiovascular outcomes such as increased blood pressure [19,20,21] and decreased femoral artery vasodilation and increased circulating endothelin-1 [22]. However, there are no previous studies exclusively targeting responses of older adults across a range of physiological systems. Indirect evidence for the adverse effects of sitting on musculoskeletal function has been shown in a previous study of older women with a mean age of 78 years, in whom sitting for only 45 minutes in a cool environment (15 degrees Celsius), was sufficient to cause reductions in muscle power (4-5%), sit-to-stand performance velocity (9%), gait speed and maximum quadriceps strength [23]. It is unclear whether sitting (and consequent muscle inactivity) in normal temperature environments would lead to reduced power in older people but clearly, muscle temperature has a powerful influence on musculoskeletal health via a reduction of muscle power output.

These data highlight the relative lack of knowledge of the adverse health outcomes of sedentariness in older adults and the mechanisms underlying them, with most of the evidence deriving from cross-sectional studies and very limited evidence from interventional study designs incorporating dose response elements studies. Addressing these knowledge gaps would allow the characterisation of sitting time in older adults in terms of acute functional and cognitive effects. Further, the knowledge gained would enable the development of interventions targeted at interrupting sitting time at specific time points, alongside generation of specific advice to older people, although this first requires the elucidation of dose-response relationships between sitting and health outcomes to be defined during carefully controlled studies. Therefore, the aim of this study was to determine the acute effects of varying durations of continuous sitting on physiological, psychological and cognitive function in adults aged 70 years or older to gain insights into the physiological mechanisms underlying the adverse effects of prolonged periods of sitting time. We hypothesised that progressively increasing sitting time would be associated with concomitant reductions in function, with lower limb power output selected as the primary outcome measure, due to its functional relevance and susceptibility to the influence of inactivity.

## Methods

The study was approved by the NHS West Midlands-South Birmingham Research Ethics Committee no. 15/WM/0260 and was registered on Clinical Trials.gov NCT026059801 and the NIHR Clinical Research Network Portfolio 19665. Participants provided informed written consent (consent form approved by Ethics Committee as above) on their first familiarisation visit in the presence of the lead researcher SA-B and a Research Nurse. Recruitment for this study was over the following period: 18/03/16 – 26/11/18.

### Participant identification, selection and recruitment

Healthy participants were identified via the Birmingham 1000 elders volunteer database held by the University of Birmingham, supported housing facilities and local older people’s clubs. Health exclusion criteria were applied to the responses to a health questionnaire and were based on those previously published for studies defining ‘medically stable’ older adults [24] (supplementary file 1). These criteria enabled the inclusion of those who had well managed long-term conditions. Participants were also recruited through newspaper and social media advertisements. Inclusion criteria were aged 70 years or older and ambulatory with or without walking aids.

### Procedures

Participants attended the Wellcome Trust Clinical Research Facility (WTCRF), (via taxi transport if required) on four occasions approximately one-week apart between 12 noon and 14:00. This facility contained temperature-controlled rooms which were set at 20°C, which is the World Health Organisation (WHO) minimum recommended indoor temperature for ‘very old’ people [25], and to minimise potential confounders such as seasonal temperature changes during the study. The first visit was a familiarisation visit, which entailed a screening assessment of cognitive function (Mini Mental State Examination, MMSE; [26]) and tissue viability (Waterlow score, [27]), with subsequent visits (2, 3, 4) to complete periods of sitting time of 1 h, 2 h and 4 h in a randomised cross-over design. A statistician generated a randomised order of sitting time for all participant visits. The sitting periods corresponded to a brief control period of sitting (1h) and classifications of ‘moderately sedentary’ (2-4 h.day-1), and ‘sedentary’ (>4h.day-1) [28]. Participants were studied on all three occasions at the same time of day (starting approximately 13:00). They were given lunch on each occasion at least 1h prior to testing. The skin was inspected prior to the sitting period, to reassess tissue viability and participants were encouraged to void urine before starting each bout of sitting time. To standardise across sessions, a standard hospital armchair containing a gel filling was used for the periods of sitting time and participants were asked to wear comfortable clothing and the same garments at each visit. They were permitted to watch television or read but they were not allowed to sleep. Participants were transferred to a toilet (ensuite) using a wheelchair if they needed to use a toilet during the period of sitting time (and this was recorded).

### Primary, secondary and exploratory outcome measures

All outcome measures were made before and after each period of sitting time, with the exception of tissue viability [27], which was measured only before each period of sitting time. The primary outcome was lower limb explosive power (LLEP) measured using the Nottingham Power Rig [29]. Secondary outcomes were timed chair rise (5-repetition Sit To Stand) [30,31]; Timed Up and Go (as a marker of falls risk; [32]); blood pressure (to determine effects on postural hypotension); perception of musculoskeletal comfort/pain (VAS) [33], vitality (Subjective Vitality Scale) [34]; mood (Profile of Mood States (POMS) [35]; acute tests of cognitive function including the Alice Heim 4 [36,37,38], Simon Task [39,40], Digit Symbol [41] and Trail Making Test B [42]. Venous blood samples were taken before and after each period of sitting time for serum inflammatory markers IL-1β, IL-6, IL-8, TNF-α and the anti-inflammatory cytokine IL-10. These cytokines were measured using a commercially available Bio-Plex Pro Human Screening Panel Assay kit according to the manufacturer’s instructions (#17004991; 5-plex; Bio-Rad, UK). Analysis was completed using BioRad Bio-Plex Manager (v 6.1). Samples of saliva required for analysis of cortisol via ELISA (IBL Hamburg, Germany), were taken at the start of each sitting session, 1 h, 2 h and 4 h later and were analysed as previously reported [43]. When the 1 h and 2 h periods of sitting time were over, participants were asked to remain in the WTCRF, to measure salivary cortisol at 4 h (to separate natural diurnal variation from the effects of sitting). If participants were unable to wait, the salivary cortisol sample was provided outside the facility at 4 h and returned approximately 1-3 hours later.

Participants were also asked open-ended questions about how they felt during and after the period of sitting, with a particular focus on feelings of mood, stiffness and pain. In a subgroup of participants (n=14), bladder volume was measured digitally before and after each sitting period using a non-invasive automated technology (Biocon 700 Cubescan, de Smit Medical Systems, UK), to investigate any association between changes in bladder volume and blood pressure during sitting, particularly after 4 h. If any participant required the toilet, we measured bladder volume just before and after voiding the bladder; however, this was not done if the participant indicated any urgency.

### Sample size

We aimed to recruit n=130 older men (n=50) and women (n=80) aged 70 y or older. The study was powered for a 10% change in the mean value of the primary outcome variable, LLEP. We have extensive reference data for older men and women up to and including the ninth decade [44] (sample sizes were calculated separately for men and women) and given that LLEP declines at an average of 1.5% per annum across this age range [45]. A decline of 10% LLEP would represent a loss of 7 ‘mobility years’. Thus, recruitment of n=130 participants would provide 90% power at a significance level of 5% including an anticipated drop-out rate of 20% to detect a 10% reduction in LLEP over an acute bout.

### Data Analysis

All data were analysed using SPSS (v 23) using two-way repeated measures ANOVA with post-hoc comparisons. For each outcome, a 3 session (1 hr, 2 hr, 4 hr) x 2-time (pre-, post-) repeated measures ANOVA with pre-specified Least Significant Difference post-hoc comparisons analysis on the post-session values was undertaken to determine between session (condition) differences by comparing values post-1 h, 2h and 4 h of sitting.

## Results

We approached n=139 older adults and consented n=70. Of these, n=65 participants were randomised to the sitting sessions, mean (SD) age 78 (7) years, range 70-99 years, n=48 female/17 male (see Fig 1: CONSORT diagram). A total of 57 participants completed all 3 sitting times, n=65 completed 1 h, n=62 completed 2 h and n=62 completed 4 h sitting. 57 participants completed all three sitting sessions 1 h, 2 h, and 4 h. Participant characteristics are shown in Table 1.

**Figure 1:**
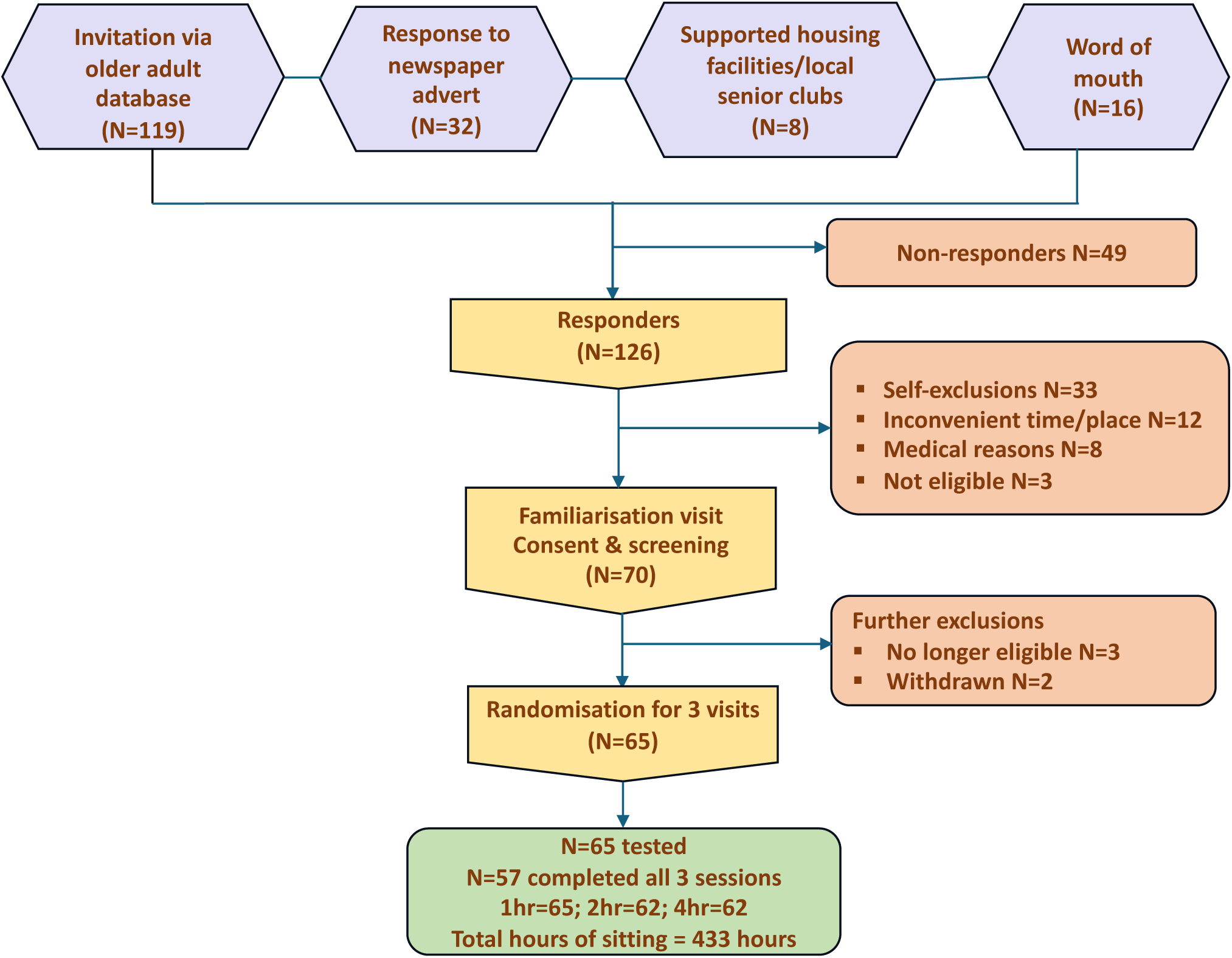
ACUSIT CONSORT diagram illustrating participant flow through the study. Self-exclusions involved participants providing personal reasons (change in circumstances; refusals; self-perceived weakness). Medical reasons included participants with recently diagnosed conditions, individuals with booked surgeries and ongoing medical investigations and appointments.

**Table 1:**
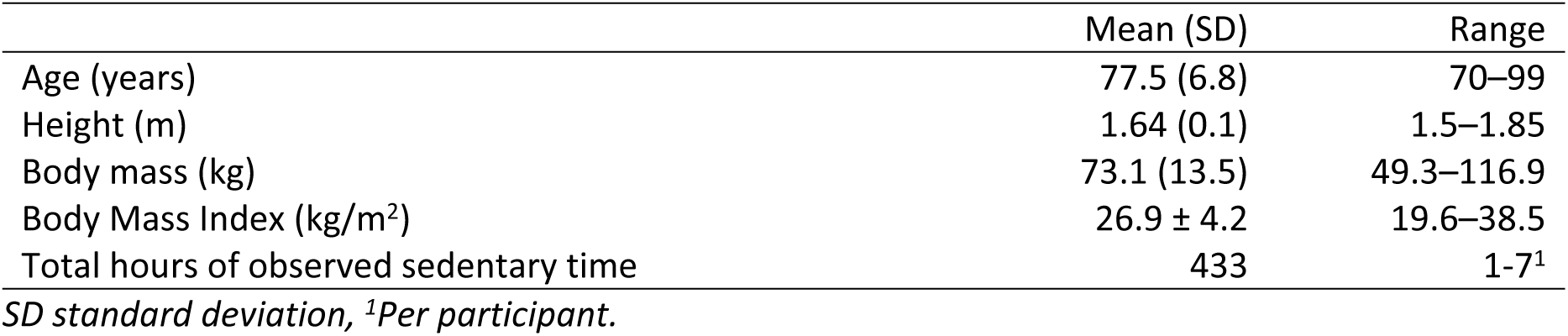
Participant characteristics.

|  | Mean (SD) | Range |
| --- | --- | --- |
| Age (years) | 77.5 (6.8) | 70–99 |
| Height (m) | 1.64 (0.1) | 1.5–1.85 |
| Body mass (kg) | 73.1 (13.5) | 49.3–116.9 |
| Body Mass Index (kg/m<sup>2</sup>) | 26.9 ± 4.2 | 19.6–38.5 |
| Total hours of observed sedentary time | 433 | 1–7<sup>1</sup> |

*SD standard deviation, <sup>1</sup>Per participant.*

### Physical function

A two-way repeated measures ANOVA (3 sessions x 2 times) for LLEP [F(2,54)=1.72, p=0.19, n^2^=.060], TUG [F(2,54)=0.96, p=0.39, n^2^=.034] and Timed Chair Rise [F(2,54)=0.16, p=0.85, n^2^=.006] showed that there were no significant session x time interaction effects for these three measures (Table 2). There were also no significant main effects of time, except for TUG (p=0.01) which worsened (i.e., time increased) across all sessions.

**Table 2:** Acute effect of prolonged sitting on physical function.

| Functional Measure | Mean ± SD (n=56) |  |  | p-value |
| --- | --- | --- | --- | --- |
|  | 1 h | 2 h | 4 h |  |
| **LLEP (Watt/kg)** |  |  |  |  |
| *Pre* | 1.44 ± 0.6 | 1.42 ± 0.7 | 1.47 ± 0.7 |  |
| *Post* | 1.46 ± 0.6 | 1.44 ± 0.6 | 1.44 ± 0.6 | 0.93<sup>1</sup> |
| *Difference* | 0.02 ± 0.2 | 0.02 ± 0.3 | -0.03 ± 0.2 | 0.84<sup>2</sup> |
| **Timed Up & Go (s)** |  |  |  |  |
| *Pre* | 7.47 ± 1.9 | 7.53 ± 2.0 | 7.48 ± 1.8 |  |
| *Post* | 7.55 ± 1.9 | 7.59 ± 2.0 | 7.64 ± 1.9 | **0.01<sup>1</sup>** |
| *Difference* | 0.07 ± 0.5 | 0.07 ± 0.5 | 0.16 ± 0.4 | 0.78<sup>2</sup> |
| **Timed Chair Rise (s)** |  |  |  |  |
| *Pre* | 11.89 ± 4.1 | 11.86 ± 4.3 | 11.95 ± 4.7 |  |
| *Post* | 12.07 ± 4.6 | 11.86 ± 4.1 | 12.06 ± 4.2 | 0.46<sup>1</sup> |
| *Difference* | 0.17 ± 1.66 | 0.01 ± 1.03 | 0.10 ± 1.9 | 0.69<sup>2</sup> |

*SD-standard deviation, LLEP-lower limb explosive power, Difference (post- minus pre-). Higher TUG and chair rise times indicate worse mobility. <sup>1</sup> indicates main effect of time; <sup>2</sup> indicates main effect of session*

### Cognitive function

Repeated measures ANOVA (3 sessions x 2 times) for Trail Maker B [F(2,57)=1.92, p=0.16, n^2^=0.063], Simon Task [F(2,54)=0.46, p=0.64, n^2^=0.017], Digit Symbol Substitution (DSS) correct scores [F(2,54)=0.11, p=0.90, n^2^=0.004], the percentage of accurate DSS scores [F(2,54=0.84, p=0.44, n^2^=0.03], Alice Heim 1 [F (2,56) =2.51, p=0.09, n^2^=0.082], and Alice Heim 2 [F (2,56) =2.15, p=0.13, n^2^=0.071] showed no significant session x time interaction effect for all four measures (Table 3). There were, however, significant main effects of time for DSS correct (p<0.001) and percentage DSS (p<0.001) which showed participants recorded better responses post sitting across all sessions. A significant difference was observed for the post-session scores on the Alice Heim 2 between 1 h and 4 h sessions (p=0.04) such that the post-session score was improved after 1 h compared to after 4 h sitting.

**Table 3:**
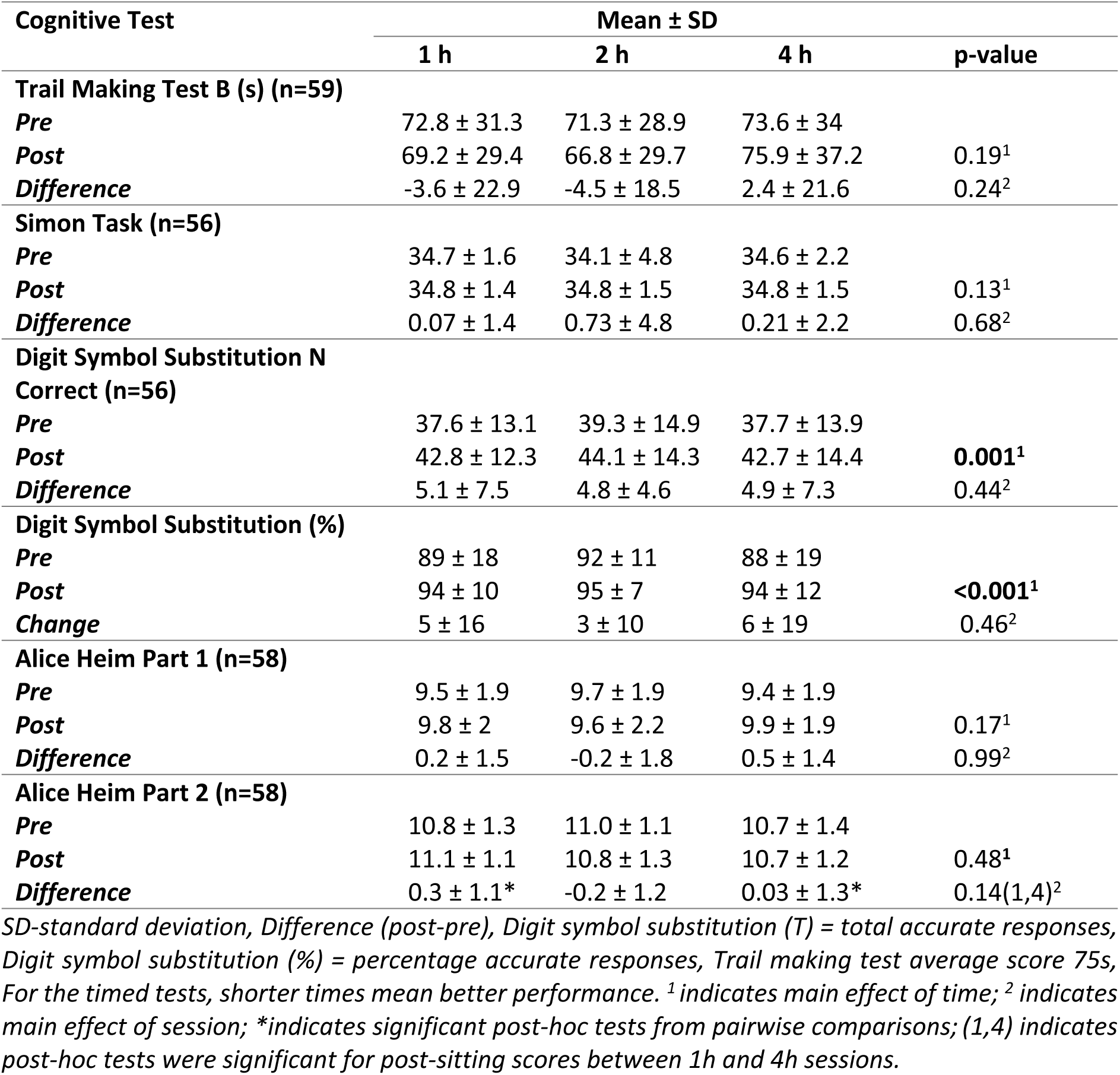
Acute effect of prolonged sitting on cognitive function.

### Psychological function

#### Vitality

Repeated measures ANOVA (3 sessions x 2 times) for vitality showed that there was a significant interaction effect, [F(2,57)=4.20, p=0.02, n^2^=0.128]. There were also significant main effects of time (p<0.001) and session (p=0.003) showing a decline across all sessions but with a dose-response showing worsening with longer sessions of sitting. Post-hoc tests showed significant differences between the post-sitting values for the 1h and 4h sessions (p<0.001) as well as between the 2 h and 4 h sessions (p=0.01) but not between 1 h and 2 h post-sitting (p=0.53) [Table 4].

**Table 4.** Effect of prolonged sitting time on vitality and profile of mood subscales.

| Psychological domain | Mean (SD) (n=58) |  |  | p-value |
| --- | --- | --- | --- | --- |
|  | 1 h | 2 h | 4 h |  |
| **Vitality (n=59)** |  |  |  |  |
| *Pre* | 29 ± 6 | 29 ± 8 | 29 ± 7 |  |
| *Post* | 27 ± 7 | 27 ± 8 | 24 ± 8 | **<0.001<sup>1</sup>** |
| *Difference* | -2 ± 4* | -2 ± 5* | -5 ± 7* | **0.003<sup>2</sup> (1,4; 2,4)** |
| **Tension** |  |  |  |  |
| *Pre* | 0.24 ± 0.5 | 0.19 ± 0.5 | 0.52 ± 1.1 |  |
| *Post* | 0.19 ± 0.5 | 0.19 ± 0.6 | 0.17 ± 0.7 | 0.13<sup>1</sup> |
| *Difference* | -0.05 ± 0.6 | 0.00 ± 0.7 | -0.34 ± 1.3 | 0.16<sup>2</sup> |
| **Depression** |  |  |  |  |
| *Pre* | 0.79 ± 2.6 | 0.71 ± 2.1 | 0.64 ± 2.3 |  |
| *Post* | 0.34 ± 0.8 | 0.43 ± 1.2 | 0.33 ± 1.0 | 0.18<sup>1</sup> |
| *Difference* | -0.45 ± 2.7 | -0.28 ± 1.8 | -0.31 ± 2.4 | 0.83<sup>2</sup> |
| **Anger** |  |  |  |  |
| *Pre* | 1.41 ± 4.7 | 1.28 ± 4.1 | 1.40 ± 5.2 |  |
| *Post* | 0.33 ± 0.8 | 0.59 ± 2.1 | 0.38 ± 0.9 | 0.07<sup>1</sup> |
| *Difference* | -1.09 ± 4.3 | -0.69 ± 3.8 | -1.02 ± 4.8 | 0.89<sup>2</sup> |
| **Fatigue** |  |  |  |  |
| *Pre* | 1.10 ± 2.0 | 1.29 ± 1.9 | 1.19 ± 1.8 |  |
| *Post* | 1.07 ± 1.4 | 1.48 ± 2.1 | 2.07 ± 2.7 | 0.16<sup>1</sup> |
| *Difference* | -0.03 ± 1.8* | 0.19 ± 2.2 | 0.88 ± 3.2* | **0.03<sup>2</sup> (1,4)** |
| **Confusion** |  |  |  |  |
| *Pre* | 0.22 ± 0.5 | 0.22 ± 0.6 | 0.21 ± 0.6 |  |
| *Post* | 0.17 ± 0.5 | 0.29 ± 0.6 | 0.22 ± 0.6 | 0.82<sup>1</sup> |
| *Difference* | -0.05 ± 0.6 | 0.07 ± 0.6 | 0.02 ± 0.6 | 0.55<sup>2</sup> |
| **Vigour** |  |  |  |  |
| *Pre* | 7.03 ± 2.5 | 6.76 ± 2.6 | 6.74 ± 2.6 |  |
| *Post* | 6.21 ± 2.6 | 5.66 ± 3.0 | 5.50 ± 2.8 | **<0.001<sup>1</sup>** |
| *Difference* | -0.83 ± 1.9* | -1.10 ± 2.1 | -1.24 ± 2.5* | **0.052<sup>2</sup> (1,4)** |
| **Friendliness** |  |  |  |  |
| *Pre* | 7.91 ± 2.2 | 8.09 ± 2.3 | 8.09 ± 2.2 |  |
| *Post* | 7.74 ± 2.5 | 7.50 ± 2.6 | 7.48 ± 2.7 | **0.03<sup>1</sup>** |
| *Difference* | -0.17 ± 1.9 | -0.59 ± 2.3 | -0.60 ± 2.4 | 0.98<sup>2</sup> |

(SD) standard deviation, <sup>1</sup> indicates main effect of time; <sup>2</sup> indicates main effect of session; \*indicates significant post-hoc tests from pairwise comparisons; (1,4) indicates post-hoc tests were significant for post-sitting scores between 1 h and 4 h sessions and (2,4) indicates post-hoc tests were significant for post-sitting scores between 2 h and 4 h sessions.

#### POMS Subscales

The repeated measures ANOVA for POMS subscales showed there were no significant interaction effects for these measures: Tension [F(2,56)=2.07, p=0.14, n^2^=0.069], Depression, [F(2,56)=0.21, p=0.81, n^2^=0.007], Anger, [F(2,56)= 0.60, p=0.55, n^2^=0.021], Fatigue [F(2,56)=2.55, p=0.09, n^2^=0.083], Confusion, [F(2,56)=0.59, p=0.56, n^2^=0.021], Vigour [F(2,56)=0.67, p=0.52, n^2^=0.023], and Friendliness, [F(2,56)=0.97, p=0.39, n^2^=0.033]. There were significant main effects of time for Vigour (p<0.001) and Friendliness (p=0.03) showing a decline across all sessions. Additionally, Vigour showed a marginal main effect of session (p=0.052), where the 1 h and 4 h sessions differed in the size of the within-session change, also indicated in the post-hoc comparisons where post-sitting Vigour significantly differed (p=0.02) between 1 h and 4 h. For Fatigue there was a significant main effect of session (p=0.03) where the change in the 1 h session differed significantly from that in the 2 h and 4 h; and post-hoc tests showed that post-sitting values were significantly different between 1 h and 4 h sessions (p=0.01) being worst after 4 h sitting [Table 4 and Figure 2].

### Physiological measures

#### Blood pressure

Blood pressure increased after each period of sitting time in a dose-response manner. The difference in mean systolic blood pressure after 4 h of sitting time compared with 1h was 10 mmHg. Two-way repeated measures ANOVAs showed that there was a significant session x time interaction effects for SBP, [F(2,56)=23.34, p <0.001, n^2^=0.455], DBP, [F(2,56)=21.35, p <0.001, n^2^=0.433], and MAP, [F(2,56)=27.06, p <0.001, n^2^=0.49]. For SBP, DBP and MAP, post-hoc tests on the post-sitting values were significantly different between each pair of sessions; 1 h, 2 h and 4 h, all p<0.01 (Table 5, Fig 3a). Importantly, the size of change was significantly different between all sessions but notably both 2 h and 4 h sessions differed in the direction of change as well compared to the 1 h session for SBP, DBP and MAP.

**Table 5:** Acute physiological response to prolonged sitting.

| Physiological measure | Mean (SD) (n=58) |  |  | p-value |
| --- | --- | --- | --- | --- |
|  | 1 h | 2 h | 4 h |  |
| **Systolic BP (mmHg)** |  |  |  |  |
| *Pre* | 139 ± 15 | 137 ± 13 | 137 ± 14 |  |
| *Post* | 137 ± 15 | 141 ± 16 | 147 ± 15 | <0.001<sup>1</sup> |
| *Difference* | -3 ± 10* | 4 ± 10* | 10 ± 10* | <0.001<sup>2</sup>(1,2; 1,4; 2,4) |
| **Diastolic BP (mmHg)** |  |  |  |  |
| *Pre* | 74 ± 9 | 74 ± 10 | 73 ± 10 |  |
| *Post* | 73 ± 10 | 76 ± 10 | 79 ± 10 | <0.001<sup>1</sup> |
| *Difference* | -1 ± 7* | 2 ± 6* | 6 ± 6* | <0.001<sup>2</sup>(1,2; 1,4; 2,4) |
| **MAP (mmHg)** |  |  |  |  |
| *Pre* | 96 ± 10 | 95 ± 10 | 95 ± 10 |  |
| *Post* | 95 ± 11 | 97 ± 11 | 102 ± 10 | <0.001<sup>1</sup> |
| *Difference* | -1 ± 7* | 2 ± 7* | 7 ± 7* | <0.001<sup>2</sup>(1,2; 1,4; 2,4) |
| **Heart Rate (bpm) (n=56)** |  |  |  |  |
| *Pre* | 67 ± 11 | 67 ± 9 | 68 ± 11 |  |
| *Post* | 62 ± 9 | 61 ± 9 | 63 ± 10 | <0.001<sup>1</sup> |
| *Difference* | -6 ± 6 | -6 ± 5 | -5 ± 6 | 0.45<sup>2</sup> |
| **Cortisol (µg/dL)** |  |  |  |  |
| *Pre* | 0.13 ± 0.1 | 0.14 ± 0.2 | 0.13 ± 0.1 |  |
| *Post* | 0.15 ± 0.3 | 0.13 ± 0.2 | 0.08 ± 0.1 | 0.07<sup>1</sup> |
| *Post 2* | 0.09 ± 0.2 | 0.08 ± 0.1 | 0.08 ± 0.1 | 0.88<sup>#</sup> |
| *Difference* | 0.02 ± 0.2 | -0.01 ± 0.2 | -0.05 ± 0.1 | **0.02<sup>2\*</sup> (1,4)** |

(SD) standard deviation, BP blood pressure, <sup>1</sup> indicates main effect of time; <sup>2</sup> indicates main effect of session; \*indicates significant post-hoc tests from pairwise comparisons; (1,2) indicates post-hoc tests were significant for post-sitting values between 1 h and 2 h sessions; (1,4) indicates post-hoc tests were significant for post-sitting values between 1 h and 4 h sessions; (2,4) indicates post-hoc tests were significant for post-sitting values between 2 h and 4 h sessions. # indicates interaction effect using pre- and post-2 values (post-2 refers to the cortisol measurement made 4 h after the start of each session). Note: there was only one measurement post 4 h; the 'post 2' value in the table is duplicated to allow the RM ANOVA interaction effect calculation.

#### Heart rate

There was no session x time interaction effect for heart rate, [F(2,54)=0.44, p=0.65, n^2^=0.016, but the main effect of time shows it significantly declined within all sessions. Post-hoc tests on the post-sitting values showed no significant differences between sessions [Table 5].

### Cortisol Concentration

A two-way repeated measures ANOVA (3 session x 2 times) for salivary cortisol concentration showed that there was no significant interaction effect, [F(2,54) =2.81, p=0.07, n^2^=0.09]. Post-hoc tests on the post-sitting values showed no significant differences between any of the sessions. Repeated measures ANOVA (3 sessions) with post-hoc tests showed that the size of the change pre-post-session was significantly different between 1 h and 4 h only (p=0.02).

### Serum Inflammatory markers

Due to inconsistencies in data output from the Bio-Plex analysis, mainly due to concentrations being out of range, (below lower limit), repeated measures ANOVA (3 sessions x 2 times) for the interaction effect of serum inflammatory markers was not feasible. Supplementary file 2 presents calculated marker levels (where data were available) for pre- and post-sitting times.

### Bladder volume

Bladder volume data were collected in a sub-sample (n=13; 77% female) aged 76.6 (5.81) years with a mean (SD) BMI of 28.2 (4.66). Repeated measures ANOVA (3 sessions x 2 times) for bladder volume showed that there was no significant interaction effect [F(2,8)=0.95, p=0.41, n^2^=0.09]. There was a significant main effect of time (p<0.001) showing a significant increase within all sessions. Post-hoc tests showed no significant differences between the 1h, 2h and 4h session post-sitting values [Table 5]. Insufficient data were obtained in our study (n=13 participants) to undertake a meaningful correlation analysis with changes in blood pressure.

**Table 6:**
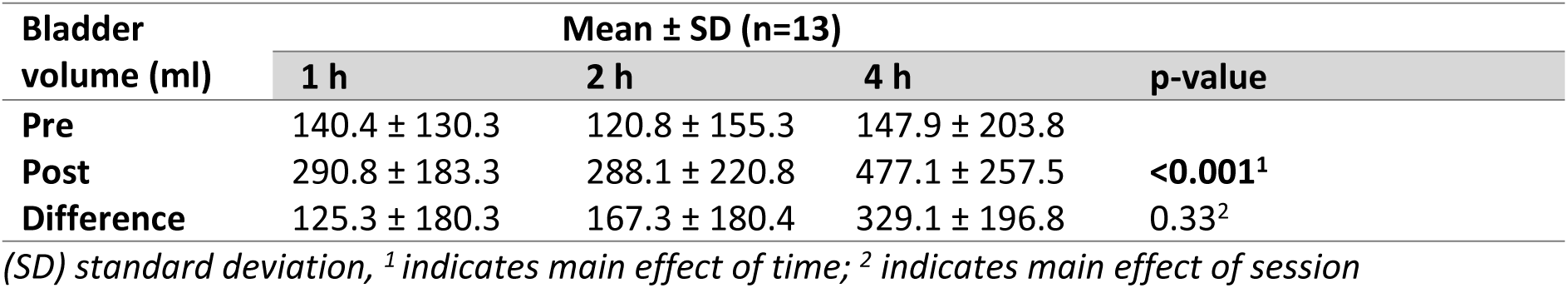
Bladder volume in response to sitting time in a study subgroup.

| Bladder volume (ml) | Mean ± SD (n=13) | | | p-value |
| --- | --- | --- | --- | --- |
|  | 1 h | 2 h | 4 h |  |
| Pre | 140.4 ± 130.3 | 120.8 ± 155.3 | 147.9 ± 203.8 | |
| Post | 290.8 ± 183.3 | 288.1 ± 220.8 | 477.1 ± 257.5 | **<0.001<sup>1</sup>** |
| Difference | 125.3 ± 180.3 | 167.3 ± 180.4 | 329.1 ± 196.8 | 0.33<sup>2</sup> |

(SD) standard deviation, <sup>1</sup> indicates main effect of time; <sup>2</sup> indicates main effect of session

## Discussion

The lack of information on dose responsiveness to acute periods of sitting across a range of outcomes, hampers the ability to determine clinically relevant ‘risk thresholds’ which could support advice to older people and those who care for them. This is a significant issue which has been highlighted in a previous international consensus and priority setting statement [46]. This is the first study to attempt to address this gap utilising a randomised dose-response design to measure responses to different periods of sitting time across a range of biological systems in a group of healthy older men and women aged 70 years or older.

The primary outcome of this study was lower limb power output, which is a functionally relevant outcome of the utmost importance to the maintenance of physical independence [44]. In this study, we hypothesised that even under controlled temperature conditions, muscle temperature would decline during prolonged sitting and that this would result in a significant decline in muscle power output. However, we found no significant differences within or between sessions. This may have been due to underpowering as we were not able to recruit the numbers of participants required in our sample size calculation. In addition, we did not measure muscle temperature directly as this would likely have affected the other measurements. Given that the study was designed to simulate a ‘real life’ situation, it was not feasible to accommodate parallel mechanistic investigations. We therefore do not know whether there was any decline in muscle temperature with up to 4h of sitting time, or whether there was a decline which was insufficient to affect power output. Although we did not observe significant changes in functional ability measured through chair rise, there was a significant effect on timed up and go performance which declined across all sessions but showed no dose-response effect. Given the ability of TUG to predict future falls and differentiate between intermediate and high-risk fallers [47,48], these data suggest that falls risk may increase after prolonged sitting even if muscle power output and strength are maintained. It is possible that a negative effect on dynamic balance and proprioception (not measured here) is more evident in a functional test requiring aspects of gait control and speed compared with the more static chair rise test [47].

We did observe significant changes in blood pressure, with the magnitude of the effect increasing between 2 h and 4 h. With respect to blood pressure, our novel data obtained from healthy adults in the eighth decade of life are consistent with a number of previous studies (reviewed in [49]), in normotensive and hypertensive middle-aged adults with overweight/obesity), which reported increases in blood pressure with prolonged sitting [19,20,21,22], with metabolic as well as vascular and autonomic mechanisms being proposed [49]. In addition, a recent systematic review and meta-analysis of blood pressure responses to acute periods of sitting time found small but significant increases in systolic blood pressure and mean arterial pressure although a non-significant increase in diastolic blood pressure [50]. That review included predominantly younger adults with only 5/22 of the eligible studies including adults with mean age of 65 years and only one in which the participants were older than 70 years [51]. Given the changes in vascular function with increasing age, it is plausible to suggest that increases in acute blood pressure in response to sitting time in older adults may be amplified compared with younger adults but since we did not include younger adults in our study, a direct comparison was not possible. Clearly, further mechanistic dose-response studies including blood pressure as a primary outcome measure, would be needed to investigate this.

The significant elevations in systolic blood pressure at the 2 h and 4 h timepoints were sufficient to render the group ‘hypertensive’, although this interpretation is a cautious one due to lack of blood pressure measurement time points during the periods of sitting time. However, our data are consistent with a recent systematic review and meta-analysis of 33 studies investigating the temporal effects of a single acute bout of sitting time (ranging from 1.5 - 10 h) on peripheral blood pressure in adults aged 19-67 years [52]. Only those studies incorporating blood pressure measurement at more than two time points were eligible for inclusion and the results showed a significant positive association between duration of sitting time and systolic, diastolic, and mean arterial blood pressure. It is known that prolonged sitting causes venous pooling in the lower limbs; indeed, previous studies of younger adults have reported decreased lower leg blood flow following 6 hours of uninterrupted sitting [53] and 4 of the studies included in [52] reported indicators of blood pooling (i.e., increases in calf circumference and decreases in gastrocnemius tissue oxygenation). A study of younger adults exposed to prolonged sitting for 3h showed a negative association between blood pooling and stroke volume which provides a potential mechanism for increased blood pressure through a decreased cardiac output, and subsequent aortic shear stress, leading to endothelial dysfunction and an increase in arterial stiffness [54]. Other potential mechanisms could also contribute to prolonged sitting induced changes in blood pressure, such as stimulation of the renin-angiotensin-aldosterone system consequent to a reduction of renal perfusion pressure [55]. Lower limb vascular dysfunction could also increase arterial stiffness and blood pressure: Previous studies using flow mediated dilation to measure lower limb arterial stiffness have reported increases with prolonged sitting [49].

Bladder distension has been shown to increase blood pressure. An earlier study in young healthy men showed an increase in sympathetic activity from recordings of the peroneal nerve. Sympathetic outflow increased in the presence of urinary urgency, and this was associated with significant increases in blood pressure [56]. In a more recent study of 172 women aged 40-60 years, significant differences in systolic and diastolic blood pressure immediately before and after voiding following at least h of urine holding (as a proxy for distension), were reported. However, there was no significant correlation between duration of urine holding and blood pressure changes [57]. Although there are no other studies to our knowledge investigating duration of sitting time with blood pressure and bladder volume measures in older adults, it is plausible to suggest that certainly after 4 h of sitting, the bladder distension we measured could be contributing to driving increases in blood pressure, alongside the other potential mechanisms suggested above.

Mood and vitality also showed some significant changes, with vigour and friendliness worsening within all sessions but vigour more so after 4 h sitting; fatigue worsening in the 2 h and 4 h sessions, more so after 4 h of sitting; and vitality declining across all sessions but being worst after 4 hr sitting. Although effects between and within sessions on mood were not consistent across all sub-scales, there was a pattern emerging for the elements of mood relating to energy/tiredness (fatigue, vigour, vitality) rather than emotion (anger, depression) or cognition (confusion). These aspects consistently showed that the post-sitting declines were greater after 4 h sitting compared to 1 h. This suggests that these relatively short amounts of sitting may not be long enough to uniformly negatively affect emotions, with the exception of friendliness, in contrast to longer periods of acute sitting such as for 6 h, where a decline in positive affect has been reported [58]. However, these sedentary bouts can contribute to energy/tiredness perceptions in line with previous studies relating sitting versus intermittent walking sessions to greater fatigue in the uninterrupted sitting condition [59,60].

In the present study, we observed a significant reduction in cortisol at 4h post start of sitting across all sessions irrespective of duration. This is consistent with what we know about cortisol levels slowly decreasing throughout the day. Samples of saliva for analysis of cortisol were taken at the start of each sitting session, and at 1 h, 2 h and 4 h later to account for the known influence of time of day. We have previously reported that different amounts of activity can relate to flatter or steeper declines in cortisol across the day [61]. Further, acute active stress can stimulate activation of the stress response system and thus an increase in cortisol [62]. However, our data suggest that the conditions employed may not have been acutely stressful in terms of stimulating cortisol responsiveness. This result is consistent with a previous study of a 2-week free-living period of sedentariness, again in younger participants in which no significant change in total urinary cortisol between conditions was reported [63]. However, the same study showed increases in negative mood (POMS-SF) compared with control (usual activity) and a significant association between mood and a stress-test induced increase in the inflammatory marker IL-6. The authors suggest that negative changes in mood could potentially interact with acute stress and result in an increase in pro-inflammatory activity [63]. We did not find any changes in inflammatory markers in our study, possibly due to limited data, but clearly the different study protocols and age groups across studies make comparisons difficult.

## Conclusion

These data show that sitting for more than 2 h may have potentially deleterious effects on blood pressure, vitality and the fatigue, vigour and friendliness sub-scales of mood in older adults. These effects appear to be somewhat dose dependent. In addition, there may be a subsequent, increased risk of falling. This study was not designed to investigate mechanisms of physiological or psychological ‘maladaptation’ to prolonged periods of sitting; however, the data provide interesting insights which pave the way for future studies to explore these phenomena in more detail.

## Data Availability

The minimal data set is available at (https://edata.bham.ac.uk/cgi/export/eprint/1780/DataCiteXML/bham_data-eprint-1780.xml)

## Perspective

The current UK and WHO physical activity guidelines state that “Older adults should break up prolonged periods of being sedentary with light activity when physically possible, or at least with standing, as this has distinct health benefits for older people” [64,65]. Our data will aid engagement with older adults about immediate individual-level benefits of reduced sitting and provide practical messages that can ensure more effective take up of an intervention among older people and effective advice for the professionals that work with older people. For example, for professionals working in residential home settings or on hospital wards/ intermediate or transition care settings (post-acute illness or surgery), the results of this research give valuable information on desirable time periods for encouraging mobilisation.

## Acknowledgements

We thank the staff of the Wellcome Trust Clinical Research Facility, University Hospitals Birmingham, and our participants for their time. We are grateful to Dr Peter Nightingale for the randomisation of participants. We are grateful to the Dunhill Medical Trust (R353/ 0514) for funding S A-B and the British Geriatrics Society for funding the measurement of bladder volume in a subgroup of participants. We thank Professor Jo Booth, Glasgow Caledonian University, for her support in provision and training in the use of the bladder scanner.

## Supplementary file 2: Acute changes in serum inflammatory markers in response to prolonged sitting

| Inflammatory marker (pg/mL) | Mean ± SD | | |
| --- | --- | --- | --- |
|  | 1h | 2h | 4h |
| **IL-1β (n=4)** |  |  |  |
| *Pre* | 0.09 ± 0.08 | | 0.09 ± 0.05 |
| *Post* | 0.05 ± 0.05 | 0.05 ± 0.02 | 0.04 ± 0.03 |
| *Change* | 0.04 ± 0.14 | | |
| **IL-6 (n=13)** |  |  |  |
| *Pre* | 0.26 ± 0.25 | 0.44 ± 0.71 | 0.24 ± 0.14 |
| *Post* | 0.22 ± 0.21 | 0.19 ± 0.17 | 0.27 ± 0.23 |
| *Change* | 0.02 ± 0.11 | | -0.03 ± 0.21 |
| **IL-8 (n=32)** |  |  |  |
| *Pre* | 2.49 ± 1.74 | 2.31 ± 2.25 | 3.03 ± 2.18 |
| *Post* | 2.19 ± 1.86 | 2.25 ± 1.83 | 2.56 ± 2.51 |
| *Change* | 0.21 ± 0.96 | 0.2 ± 0.89 | 0.31 ± 1.35 |
| **IL-10 (n=4)** |  |  |  |
| *Pre* | 1.32 ± 0.89 | 3.57 ± 3.39 | 0.85 ± 0.45 |
| *Post* | 1.25 ± 1.41 | 0.79 ± 0.29 | |
| *Change* | -0.38 ± 0.97 | | |
| **TNF-α (n=10)** |  |  |  |
| *Pre* | 2.29 ± 1.8 | 2.88 ± 2.44 | 1.84 ± 1.71 |
| *Post* | 2.27 ± 2.25 | 1.69 ± 0.91 | 2.13 ± 1.88 |
| *Change* | -0.09 ± 0.86 | 0.54 ± 0.97 | 0.21 ± 0.51 |

(SD) standard deviation; IL1- $\beta$ Interleukin 1-beta; IL-6 Interleukin 6; IL-8 Interleukin 8, IL-10 Interleukin 10; TNF $\alpha$ Tumour Necrosis Factor-alpha. Units are expressed in picograms/millilitre (pg/mL).

## Supplementary file 1: Exclusion criteria (based upon those defining ‘medically stable’ participants (Greig CA et al. Age Ageing 1994, 23: 185-925)

### 1.1.8 Exclusion Criteria

These can include:

- History of myocardial infarction within previous 2 years
- Cardiac illness: moderate/ severe aortic stenosis, acute pericarditis, acute myocarditis, aneurysm, severe angina, clinically significant valvular disease, uncontrolled dysrhythmia, claudication within the previous 10 years
- Thrombophlebitis or pulmonary embolus within the previous 2 years
- History of cerebrovascular disease (CVA or TIA) within the previous 2 years
- Acute febrile illness within the previous 3 months
- Severe airflow obstruction
- Uncontrolled metabolic disease (e.g., thyroid disease or cancer)
- Significant emotional distress, psychotic illness or depression within the previous 2 years
- Lower limb fracture sustained within the previous 2 years; upper limb fracture within the previous 6 months; non arthroscopic lower limb joint surgery within the previous 2 years
- Any reason for loss of mobility for greater than 1 week in the previous 2 months or greater than 2 weeks in the previous 6 months
- Poorly controlled atrial fibrillation
- Poor (chronic) pain control
- Resting systolic pressure >200 mmHg or resting diastolic pressure >100mmHg
- Moderate/ severe cognitive impairment (MMSE <23)
- Impaired tissue viability (defined by a Waterlow risk assessment score >15).

